# Multi-omics investigation on the prognostic and predictive factors in metastatic breast cancer using data from Phase III ribociclib clinical trials: A statistical and machine learning analysis plan

**DOI:** 10.1101/2023.08.30.23294367

**Authors:** Thibaud Coroller, Berkman Sahiner, Anup Amatya, Alexej Gossmann, Konstantinos Karagiannis, Ravi K. Samala, Luis Santana-Quintero, Nadia Solovieff, Craig Wang, Laleh Amiri-Kordestani, Qian Cao, Kenny H. Cha, Rosane Charlab Orbach, Frank H. Cross, Tingting Hu, Ruihao Huang, Jeffrey Kraft, Peter Krusche, Yutong Li, Zheng Li, Ilya Mazo, Conor Moloney, Rahul Paul, Jason Plawinski, Susan Schnakenberg, Paolo Serra, Sean Smith, Chi Song, Fei Su, Sajanth Subramaniam, Mohit Tiwari, Colin Vechery, Xin Xiong, Juan Pablo Zarate, Jonathan Ziegler, Hao Zhu, Arunava Chakravartty, Qi Liu, David Ohlssen, Nicholas Petrick, Julie A. Schneider, Mark Walderhaug, Emmanuel Zuber

## Abstract

In 2020, Novartis Pharmaceuticals Corporation and the U.S. Food and Drug Administration (FDA) started a 4-year scientific collaboration to find novel radiogenomics-based prognostic and predictive factors for HR+/HER2-metastatic breast cancer under a Research Collaboration Agreement. This manuscript aims to detail the guiding principles and methodology for this study. We include a discussion of internal and external clinical, genomics, imaging datasets, data processing workflows, and machine learning model development strategies. We also prospectively define our success criteria to ensure robust scientific outputs.

**Disclosure:** This publication reflects the views of the authors and should not be construed to represent FDA’s views or policies.

## 1 Introduction

In 2020, Novartis Pharmaceuticals Corporation and the U.S. Food and Drug Administration (FDA) started a 4-year scientific collaboration to find novel prognostic and predictive factors for Hormone receptor-positive (HR+)/ human epidermal growth factor receptor 2-negative (HER2-) metastatic breast cancer (MBC) under a Research Collaboration Agreement. This collaboration includes two main goals: (1) pilot new approaches for transferring large datasets (i.e., terabytes scale) to the FDA and (2) explore new approaches to analyze patient information (clinical, genomics and/or imaging) to predict clinical outcomes (such as progression-free survival). Additionally, we will benchmark our findings to well understood risk factors (i.e., clinically established) to ensure their value. The project will achieve these goals through analyzing data from MONALEESA2, MONALEESA3, and MONALEESA7 clinical trials and may also yield meaningful scientific and clinical findings for publication.

Machine learning (ML) has shown great potential in advancing precision medicine. As compared to traditional statistical modeling, state-of-the-art ML algorithms may offer the advantages of superior predictive performance and the ability to handle high-dimensional data (such as imaging) for prediction of treatment response. Deep learning, a subset of ML, has been successfully applied to genomics and imaging data for precision medicine and to imaging data for diagnostic applications.^1^ Radiomics is a burgeoning field that uses algorithms to extract high throughput features from medical images using either engineered methods (i.e., using pre-defined recipes) or models that are trained end-to-end. Radiomics has the potential to uncover disease features and characteristics that cannot be appreciated by the naked eye.^2–4^ By extracting features from medical images, radiomics enables us to easily use medical images in a new way to help in disease detection, characterization, staging, as well as assessment and prediction of treatment response. A related field, radiogenomics, assesses the relationship between genes and phenotypes^5,6^ by combining both radiomics and genomics information. Oncology is an ideal environment to test the capabilities of these new analytical tools, due to the relevance of imaging and genomics data for cancer patients. Extensive imaging and genomic information are routinely collected in modern oncology clinical trials to characterize and follow-up disease progression. Adding both sources of information to traditional clinical data provides a richer source of data, but also presents new challenges in data fusion.

Development of ML models that use multimodal data such as imaging, genomic and clinical domains are hindered by the lack of large datasets that are needed for modern data-driven ML techniques and by to the complexity of merging information that may represent different patient and lesion level information at different time points. In addition, there are differences in the frequency and type of data collection for patients across different clinical trials which can further complicate the data fusion task. The goal of the collaboration is to explore methods to analyze the relationship between patient’s baseline characteristics, genomics and imaging data and clinical outcome. The study will also provide insight into how retrospective data from multiple clinical trials conducted at multiple sites, with a heterogeneity of collection methods and protocols, can be harmonized for this purpose.

The project will achieve these goals through analyzing data from three Phase III clinical trials in metastatic breast cancer using ribociclib as the investigational drug, namely MONALEESA-2, MONALEESA-3, and MONALEESA-7, and we expect that our efforts to yield meaningful scientific and clinical findings for publication. This project will provide an opportunity for the FDA and industry scientists to learn and implement these novel approaches to prepare for future regulatory submissions. Key research outputs from the project will be publicly shared to encourage and promote open science in oncology.

### 1.1 Project impacts

This research project is motivated by multiple expected impacts on regulatory science. First, it provides an opportunity to investigate methods for the transfer of large datasets to the FDA, as described in Section 2.1 below. As genomics, radiomics and image analysis methods advance, it may be expected that future FDA submissions will contain datasets that are large both in terms of the number of patients and files to be transferred per patient (e.g., digital pathology whole slide images that may be multiple GB in size per patient). The experience gained from the transfer of large datasets to the FDA has the potential to decrease the review time for products to be submitted to the FDA in the future. Second, the tools being developed to analyze radiomics and genomics data may have the potential to identify characteristics or patterns to enhance patient treatment options, with applications not only in breast cancer treatment but in other studies that involve fusion of imaging, genomics and clinical data. Identification of novel biomarkers or algorithms for the prediction of a patient’s prognosis or treatment outcome, which can be used to guide precision treatment based on the individual’s unique characteristics, would benefit patients worldwide. The integration of AI/ML into precision medicine holds great promise and the FDA has a crucial role in guiding its development and application in a responsible manner. This role includes careful review and evaluation of products that use AI/ML during development to make sure that the models are reliable and properly validated. The third potential benefit of this project is its ability to inform one possible way by which novel, complex devices that combine multiple sources of information are reviewed and evaluated by the FDA. In this regard, the project is expected to:

1. provide expertise and evidence on how data from clinical trials can be leveraged to develop multimodal models for prediction of treatment response so that developers have a least burdensome path for safe and effective devices for this purpose;
2. allow for a better understanding of how multimodal data from multiple sites, acquired under different conditions, can be harmonized so that the FDA can provide better recommendations to developers for this purpose; and
3. provide hands-on experience in multimodal data fusion to better understand potential design pitfalls so that the FDA can lead developers in the right path in product reviews.

## 2 Methods

### 2.1 Large Data Transfer

The ability to transfer large datasets into the FDA environment securely was an important FDA goal for this project, as new data sources and analytic methods (such as the radiomic data set analyzed for this research project) may be included in future regulatory submissions. FDA developed the Electronic Submissions Gateway (ESG), as an Agency-wide solution for accepting electronic regulatory submissions. The FDA ESG enables the secure submission of premarket and post market regulatory information for review. As the ESG has been designed to facilitate transfer of regulatory data from sponsors to the agency, it fulfills the safety and security requirements, but, at present does not have the capacity to accommodate the large datasets required for this study. Hence, this project explored new methods to transfer terabytes of clinical, genomics and imaging data and has redeveloped a potential mechanism, which can be leveraged in the future for communication of large regulatory datasets as well as research datasets. The research team leverages cloud provider storage services in combination with the on premises high performance computing HIVE^7^ platform at FDA. The process for transferring the dataset initiated from the collaborator to an Amazon Web Service (AWS) Simple Storage Service (S3) private bucket, then granting permissions to access and download files from the bucket via Access key and Secret Access Key. The credentials (private keys) were then shared with the FDA collaborators in a secure manner. FDA has developed an application in the web accessible HIVE system, based on the AWS command line interface, that enables network resilient downloads of large datasets from AWS. The download of the collaborator dataset was completed successfully in the preproduction instance of the HIVE platform. The production environment of HIVE has the authorization to operate (ATO) and it is a FISMA moderate system that hosts regulatory big data and computation, deeming this solution an option for regulatory big data and the completion of this goal a successful precursory pilot.

### 2.2 Data Sources Overview

#### 2.2.1 Internal datasets

The internal datasets used in this project originated from three randomized, double-blind, placebo-controlled, phase III clinical trials, namely MONALEESA-2, MONALEESA-7, and MONALEESA-3 which used ribociclib as the investigational drug and led to the approval of ribociclib as a targeted therapy for metastatic breast cancer. **Table** 1 summarizes the key characteristics across the three trials.

**Table 1:** An overview of the three clinical trials used in this project.

| STUDYID | MONALEESA2 | MONALEESA7 | MONALEESA3 |
| --- | --- | --- | --- |
| Sample size<br>(treatment vs placebo arm) | 334 vs 334 | 335 vs 337 | 484 vs 242 |
| Treatment arms | Ribociclib + Letrozole<br>vs Placebo + Letrozole | Ribociclib + NSAI/Tamoxifen<br>vs Placebo + NSAI/Tamoxifen<br>Both arms additionally<br>receive goserelin | Ribociclib + Fulvestrant<br>vs Placebo + Fulvestrant |
| Patient population |  |  |  |
| Menopausal status | Postmenopausal | Pre and perimenopausal | Postmenopausal |
| Lines of therapy<br>for metastatic setting | First line | First line<br>(Prior chemotherapy allowed) | First & second line,<br>Early relapse |
| Randomization<br>stratification factor | Lung and/or liver metastasis | Lung and/or liver metastasis,<br>prior chemotherapy,<br>endocrine combination partner | Lung and/or liver metastasis,<br>line of therapy |
| Primary endpoints | Progression Free Survival (PFS)<sup>1</sup> by local investigator per RECIST 1.1 criteria,<sup>8</sup><br>with Progression Free Survival by Blinded Independent review committee (BIRC) as<br>supportive evidence |  |  |
| Secondary endpoints | Include Overall Survival (OS), Overall Response Rate<sup>†</sup>, Clinical Benefit Rate<sup>‡</sup>. |  |  |

<sup>1</sup> Progression-free survival defined as the time from the date of randomization to the date of the first documented progression or death due to any cause.
<sup>†</sup>Overall response included a complete or partial response.
<sup>‡</sup>Clinical benefit in the overall population was defined as a complete or partial response, stable disease lasting 24 weeks or more, or neither a complete response nor progressive disease lasting 24 weeks or more.

Ribociclib is an orally bioavailable, selective small molecule inhibitor of cyclin-dependent kinases 4 and 6 (CDK4/6). Ribociclib has been approved by several Health Authorities, including the FDA and the European Commission, for the treatment of women and men (U.S. only) with hormone receptor (HR)-positive, human epidermal growth factor receptor 2 (HER2)-negative advanced or metastatic breast cancer in combination either with an aromatase inhibitor or with fulvestrant as initial endocrine-based therapy or following disease progression on endocrine therapy.

The patients were enrolled from 2013 until 2016 across the three trials. The data collected up to the final PFS analysis of each study were used in this project, which corresponds to median durations of follow-up of 15.3 months, 19.2 months, and 20.4 months respectively. All three trials have met the primary objective of PFS, demonstrating statistically significant improvements in PFS in the ribociclib arm compared to the placebo arm. The studies also met secondary objective of OS with longer follow-up. However, those long-term data are initially not in the scope of this project. The datasets from these three clinical trials can be categorized into three different modalities:

- Clinical data collected from electronic Case Retrieval Forms (eCRF)
  1. Data collected only at baseline include demographics, past and current medical conditions, diagnosis and extent of cancer, and prior anti-cancer therapy.
  2. Data collected at both baseline and after patients started treatment include tumor assessments (lesion diameter and location) and patient reported outcomes at every 8 weeks during the first 18 months then every 12 weeks thereafter; laboratory assessments and physical examination every 4 weeks; and ongoing adverse events reporting.
- Genomics (and transcriptomics) data generated from tumor and plasma samples
  1. All 3 trials had a mandatory archival formalin-fixed paraffin-embedded (FFPE) tumor sample (i.e., a tumor block or slides) collected prior to the start of the trial. Tumor samples were reviewed by a central lab and were excluded if the tumor content was less than 10
  2. RNA was extracted from tumor samples and gene expression was generated for 870 genes using a custom CodeSet gene panel and the nCounter platform (both from Nanostring Technologies).
  3. Plasma samples were collected at baseline and cell-free DNA was extracted using the QIAamp Circulating Nucleic Acid Kit (Qiagen). Cell-free DNA was then sequenced on an Illumina HiSeq instrument using a targeted panel of 570 genes.
- Imaging data obtained through tumor scans saved in DICOM format
  1. CT/MRI scans of chest, abdomen and pelvis performed every 8 weeks during the first 18 months on trial and every 12 weeks thereafter
  2. With or without contrast agent depending on contraindications.
  3. Whole body bone scans at Screening with local following of lesions throughout the study (CT, MRI, or X-Ray)
  4. Annotations (2-dimensional) drawn by two independent radiologists, a polygon for target lesion and a square for non-target lesion (except for MONALEESA-2, where the annotation consisted of a line along the long diameter for target lesion and a circle for non-target lesion). Annotations were available for 40% of patients from MONALEESA-3 and MONALEESA-7 and 100% of patients from MONALEESA-2 because of the BIRC strategy of respective studies.

### 2.2.2 External datasets

In addition to the clinical data from MONALEESA trials, publicly available external cancer-related datasets are leveraged to enhance the model performance. Those datasets can be used during training (increase our sample size, enable us to create other models such as segmentation tools) or as external independent testing sets. They can also be used to annotate the biological significance of somatic mutations in cancer. They include but are not limited to datasets from the cancer imaging archive (TCIA)^9^ and COSMIC^10^ as shown in **Table** 2. Our TCIA datasets are composed of de-identified medical images of solid tumors cancer patients (head & neck, and lung). We are focusing on lung CT images for this project as this is the most common modality and organ in our internal datasets. TCIA datasets are sometimes linked to clinical information such as overall survival (OS), and thus may be used either as an independent testing set or add to our training set to increase our effective sample size. All TCIA images have been processed with the same parameters as our internal datasets (2.2).

**Table 2:** Overview of publicly available imaging datasets with their respective clinical endpoints. **OS**: Overall survival, **TTE**: Time-to-event, **CT**: Computed tomography, **MRI**: Magnetic resonance images, **RT**: Radiotherapy structure

| Dataset name | Sample size | Modalities | Annotations | Relevant targets | Location |
| --- | --- | --- | --- | --- | --- |
| LCTSC | 60 | CT | RT | None (screening cohort) | link |
| NSCLC-radiomics | 422 | CT | RT | OS, Stage, Histology | link |
| NSCLC-radiogenomics | 89 | CT | RT | Stages, Histology, Lesion diameter | link |
| HN1 | 137 | CT | RT | OS, Stage, TTE recurrences | link |
| HNSCC | 627 | CT, MRI | RT | OS, Stage | link |

Over time cancer cells accumulate mutations with many variants having unknown significance, complicating model building to predict cancer outcomes and treatment strategies. COSMIC, the Catalogue of Somatic Mutations In Cancer, provides a comprehensive resource of somatic mutations in human cancer,^10^ including the COSMIC Cancer Mutation Census (CMC), a database which combines manually curated information with cancer and non-cancer population variant frequencies to evaluate and rank somatic mutation significance.^11^

### 2.2.3 Pretrained models

The use of pretrained models will be explored for several tasks, ranging from increasing our sample size of tumors by using segmentation models to the use of domain-trained models to extract more meaningful representations than using models trained on natural images (e.g, ImageNet). For segmentation, we use a previously developed internal model from TCIA images to segment lung cancer lesions from CT images using TensorFlow 2.0.^12^ This method returns 3D masks for each predicted lesion in a scan, their probability of being cancer and their center of mass coordinates. This model can be used to generate annotations from patients that did not go under central review (thus missing radiologist segmentation). It would allow us to increase our effective imaging samples. Those new samples could be either used for training or testing depending on which dataset they were assigned to. We will also investigate publicly available models from top academic centers.^13–15^

## 3 Data handling and pre-processing

Prior to any modelling attempt, several pre-processing steps are involved to ensure that the patients’ privacy remain intact, data is of sufficient quality and is in a format that is easy to handle by the models. In this section, we describe the anonymization done to the datasets in each modality and the subsequent pre-processing steps for each data modality.

### 3.1 Anonymization

A re-identification risk assessment was performed for the anonymization of the datasets from clinical trials MONALEESA-2, MONALEESA-3 and MONALEESA-7. A separate risk assessment was performed on the data for each study individually, while a common anonymization strategy was applied across all three trials.

For both the imaging and clinical datasets (including genomics) containing patient information, the methodology used for risk measurement and anonymization satisfied contemporary criteria for anonymization methodologies.^16,17^ These criteria were derived from existing standards published by regulators, government agencies, and professional groups.^18–22^ Three different plausible attacks on the data were considered as part of the assessments, these include 1) deliberate attempt; 2) inadvertent access and 3) data breach. In order to keep the probability of re-identification for each of the plausible attacks below a certain threshold, key anonymization steps included masking randomization numbers and unique IDs, generalizing continuous and date variables (such as age, date of initial cancer diagnosis) into categories, suppressing categorical variables and rare categories (such country, ethnicity, prior medical history), shifting date of events (such as laboratory assessment date, adverse event date) while keeping the consistency of dates across different events. After the anonymization steps for clinical, genomics and imaging datasets, the anonymized data are ingested into a computing platform hosted by Novartis, accessible by both Novartis and FDA teams.

### 3.2 Patient criteria selection rules

After anonymization, we subsequently labelled patients from the three clinical trials into different categories based on their data modality availability:

1. **Clinical only (C)**: must have baseline and post-baseline data collected from eCRF.
2. **Genomics (G) &** transcriptomics: must have tumor circulating DNA and NanoString information available
3. **Imaging (I)**: must have baseline and week 8 images, both with annotations (metadata indicating tumor location(s) on the image)
4. **Super (S)**: all the above

Clinical information (C) was available for all patients. We grouped patients that have genomics data but not imaging data as defined above into category (G). In a similar manner, patients that have imaging data but not genomics data as defined above were grouped into category (I). Patients satisfying both imaging (I) and genomics (G) were labeled as super (S) samples. Super samples will enable us to compare our different omics models in a consistent manner as well as combining the models in a learnable fashion. We show in **Table** 3 and **Figure** 1 the patient distribution based on our classification system.

**Table 3:** Overview of the data caterogy availability across all trials and the pooled dataset. All samples have clinical information. Data sources indicates if Genomics (G), Imaging (I) or both (S) are available in completement of clinical data source.

| STUDYID | C | G | I | S | Total |
| --- | --- | --- | --- | --- | --- |
| MONALEESA2 | 83 | 40 | 314 | 231 | 668 |
| MONALEESA3 | 196 | 279 | 100 | 151 | 726 |
| MONALEESA7 | 288 | 159 | 125 | 100 | 672 |
| Total | 567 | 478 | 539 | 482 | 2066 |

**Figure 1:**
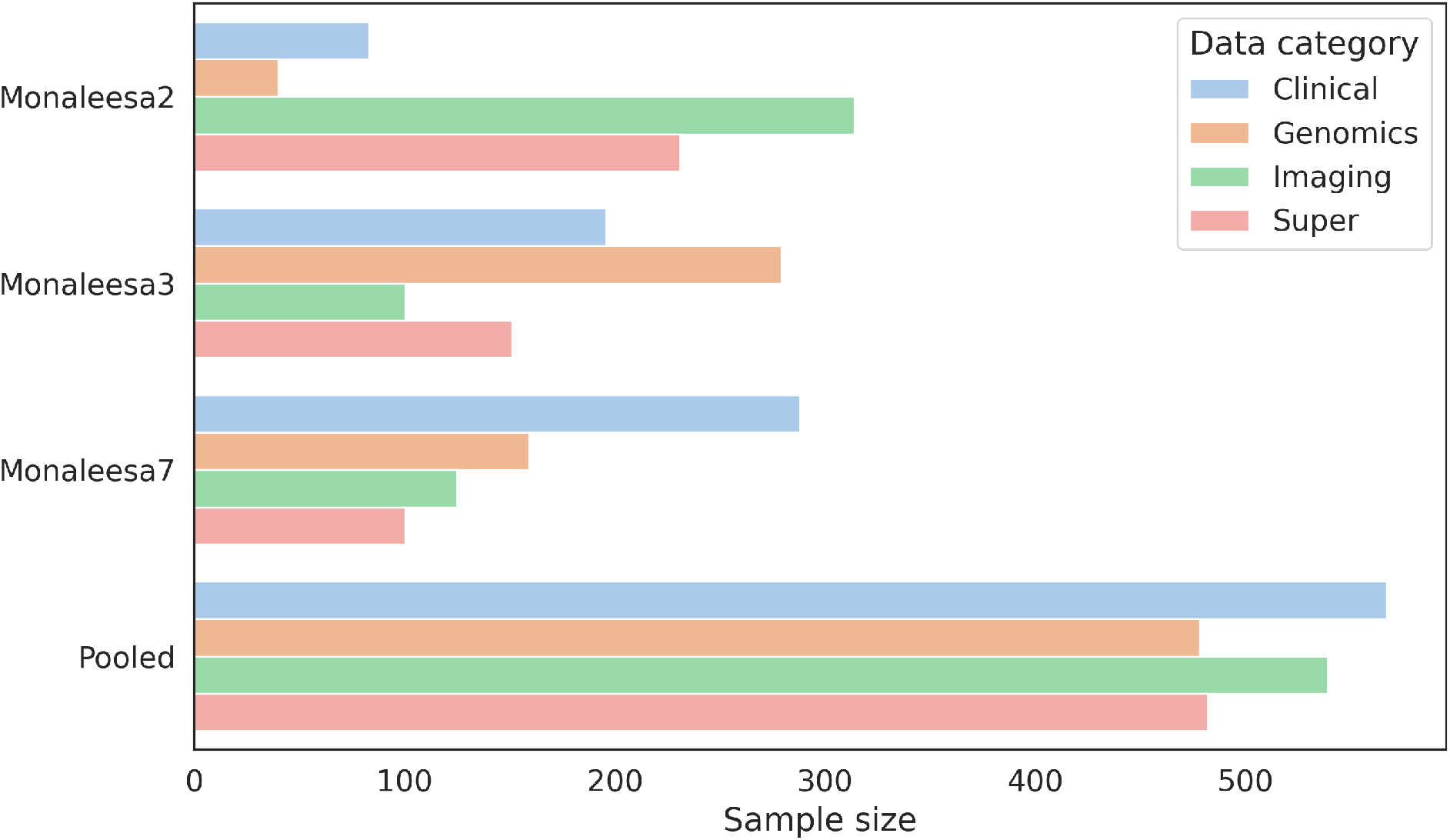
Data availability distribution per trial. Super samples are defined as patients with clinical, genomics and imaging data available.

### 3.3 Clinical

Clinical data collected from eCRF are used to directly address the key objectives in the clinical trials. They are further processed following [CDISC](https://www.cdisc.org/standards) standards during clinical trial reporting activities and for the purpose of regulatory submission of the trial results. Therefore, these data often do not require further processing. In addition, several baseline clinical attributes based on prior therapy, extent of cancer and laboratory assessments are known to have strong prognostic value in disease progression related outcomes. For these reasons, a pooled dataset with one row per patient containing baseline demographics, prior therapy, extent of cancer and laboratory assessments was merged from multiple data files for the convenience of modelling.

### 3.4 Genomics & Transcriptomics

Several cleaning and filtering steps were implemented for the gene expression and ctDNA datasets prior to model building. Gene expression: Samples with more than half of house-keeping genes expressed below background noise were removed. For patients with multiple biopsies, the most recent biopsy was retained. To normalize the raw counts, positive control and housekeeping gene normalization was performed. CtDNA: Single-nucleotide variants and indels were called using MuTect^23^ and Pindel,^24^ respectively. Copy number alterations were called, accounting for the ploidy and purity of the sample, using PureCN.^25^ Amplifications were defined as ≥6 copies and ≥7 copies for focal and non-focal events, respectively, and deletions were defined as 0 copies. Several steps were implemented to remove germline variations and sequencing artifacts:

- Comparison with ExAC database, dbSNP database and an internal database (Novartis Institutes for BioMedical Research) of normal circulating free DNA samples from healthy individuals without cancer.
- SNVs and indels with a variant allele fraction < 1% were removed unless the alteration was a hotspot in the COSMIC database
- Given that matched normal sample was not assayed, SNVs and indels with a variant allele fraction > 40% were removed unless the alteration was reported in the COSMIC database
- Samples that were collected after first exposure to study treatment were removed from downstream analysis.

Additionally, all analyses will focus on non-synonymous alterations. As part of the modeling efforts, various approaches of feature engineering will be considered to account for the functional impact of each alteration and/or whether the alteration is reported in the COSMIC database.

### 3.5 Imaging

We ingested medical images using a python package developed by Novartis called nip (Novartis Imaging package). It provides a solution to index images, merge against clinical data and export images into NumPy^26^ arrays alongside their annotation (when available). Nip is built using PyDicom^27^ and simpleITK^28^ to deal with image ingestion, metadata parsing and pre-processing (e.g., voxel resampling, normalization) As part of ingestion, the information from the DICOM^29^ header and the imaging data were separated and stored in different data formats and provide patient- and lesion-level link between the imaging data and the clinical data. These steps were motivated to train machine learning models that are based on traditional machine learning approaches that use radiomics feature extraction and deep-learning machine learning approaches that may use image patches that are based on 2-dimensional or 3-dimensional (3D) region-of-interest (ROI) or the entire slice or a 3D volume-of-interest. We considered the following to keep an image in the investigation, the image must:

1. Be a computed tomography (CT) image
2. Be from either the chest, lung, or abdominal region
3. Have been successfully ingested into a tensor without error (either technical or due to corrupted data)
4. Not have artifacts (i.e., motion, voxel intensity inaccuracy and heterogeneity) throughout the patient image

We also considered other important criteria, without strict rules within the collaboration:

- Differences in image quality: Variability in image acquisition could be mitigated by harmonization, either in the imaging domain (e.g., resampling, HU normalization, GAN-based^30^) or in the feature domain(e.g. ComBat^31–33^). It is known that radiomics features are highly susceptible to differences in image quality such as blur and noise texture. A consistent pre-processing or harmonization pipeline is key to obtaining predictive features.
- Acquisition with imaging contrast: Patient scanned without contrast agents may be dropped from specific analysis. The percentage of non-contrast studies over the three trials was about 10%.
- Lesion localization: For both radiomics and deep learning models, it may be necessary to consider only patches of the image associated with target and non-target lesions. We will use radiologist annotations of corresponding lesions over time for delta radiomics analysis and other feature extraction pipelines.

## 4 Data splitting strategy

Our three internal trials have slightly different patient population and distribution of baseline characteristics. Thus, we decided to define our training and testing set using stratified random sampling by data availability and study as seen on **Table** 4. External datasets will be used as is.

**Table 4:**
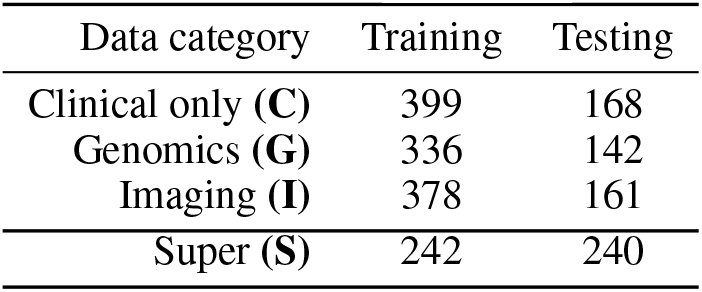
Data strategy sample size.

| Data category | Training | Testing |
| --- | --- | --- |
| Clinical only (C) | 399 | 168 |
| Genomics (G) | 336 | 142 |
| Imaging (I) | 378 | 161 |
| Super (S) | 242 | 240 |

After pre-processing, data was partitioned by data source (C: clinical, G: genomics & transcriptomics, I: imaging and S: supers) to create our training and testing sets as seen in **Figure** 2. This was done to ensure that our training and testing data would both have representative samples from each data domain by using a random stratified split with data source:

**Figure 2:**
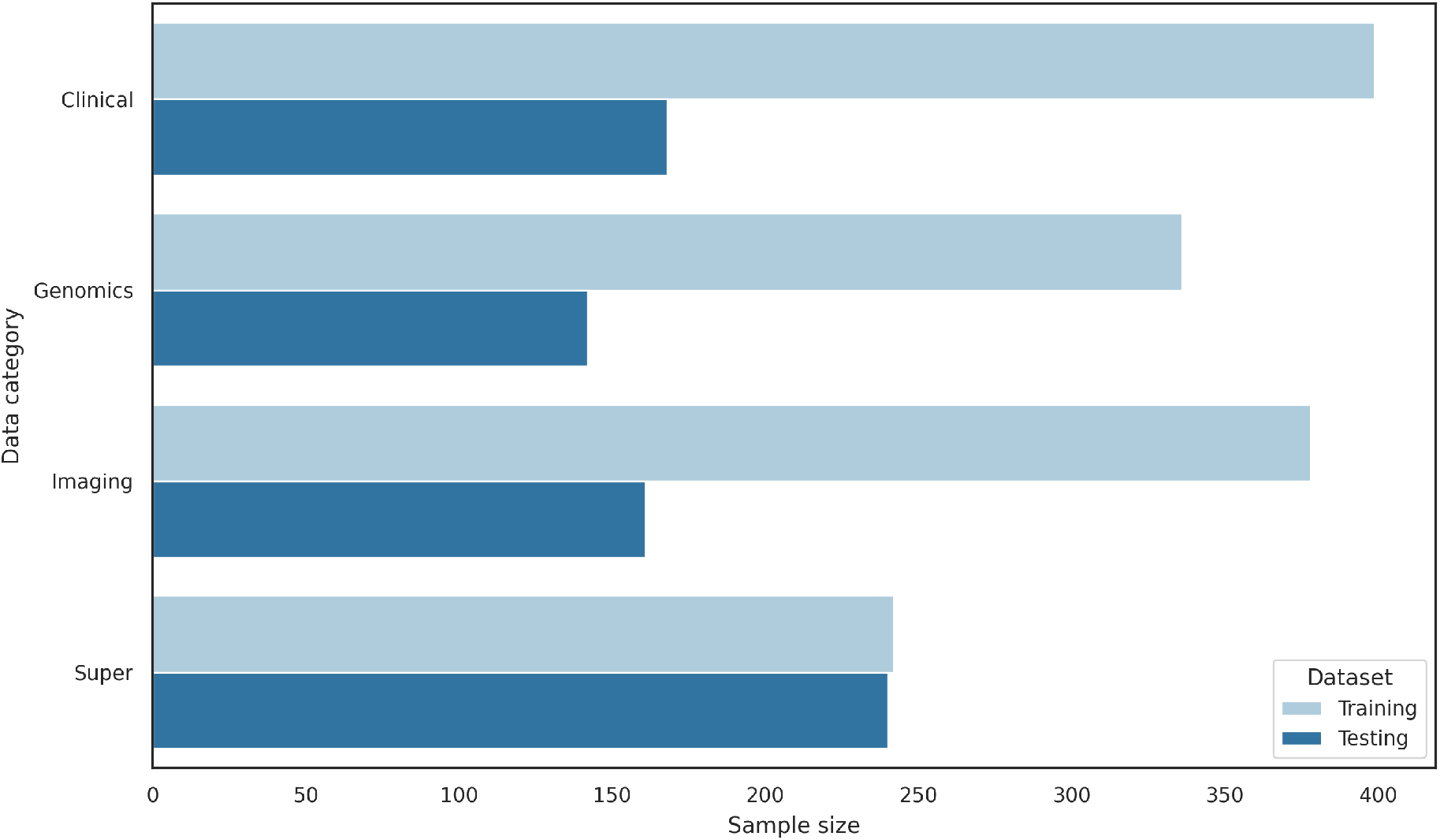
Our data splitting strategy uses a 70/30 stratified random split for the clinical, genomics and imaging data. We used a 50/50 stratified random split for super samples to ensure enough sample for model evaluation.

- 70%/30% random stratified split was used for Clinical, Genomics and Imaging domains
- 50%/50% random stratified split was used for the Super domain to leave sufficient sample size for final testing

## 5 Modelling strategies

### 5.1 Data missingness

When working with multimodal data, dealing with data missingness is key to keeping as many samples as possible (even ones with partial information) and robustly model our endpoint. This is crucial especially when working with early fusion (see section below) where the model is trained from scratch using all overlapping samples.

- **In clinical data**, the specific variables collected by the three clinical trials do not completely overlap. Variables which are only collected in one or two out of the three clinical trials will be excluded from training since they are likely to have lower importance and it is not feasible to impute them for an entire study. Additionally, there are often occasions where patients miss study visits or a subset of study assessments at certain visits. Those variables tend to have a lower missingness and can be imputed based on the data collected, for example imputing using the median or mode of non-missing values or using nearest-neighbor approaches.
- **In genomics data**, missing data generally implies the entire sample was not assayed. For example, a patient may have had a low-quality tumor sample and thus has missing values for all genes for transcriptomics. Therefore, during data pre-processing, such a sample could be excluded. One possible multimodal modelling strategy is to build transcriptomics only or genomics only models separately to maximize the number of samples with available data and then leverage late fusion methods to combine the model.
- **In imaging data**, missing data strategy could result in using an empty input (zero-filled tensor) when an image is not available. This is especially important when looking in post-baseline analysis (e.g., all time steps).^34^ Deep learning models will handle those empty tensors. If using such approach, during training we plan to randomly dropout all images (empty and others) to prevent the model from learning correlation between missing data and patient status (i.e., a patient missing time steps could be sign of early death, known as informative censoring). Any imputations on the missing values will be done during the model building stage which can be model-dependent hence we do not impose a pre-defined imputation method.

### 5.2 Classification

To have an early evaluation of patients’ clinical outcome, different criteria have been developed to classify patient’s tumor response.^8^ A patient is classified to have a response (also known as objective response) if the patient has a best overall response of complete response (CR) or partial response (PR). Other best overall response categories include stable disease (SD), progressive disease (PR) and not evaluable. Typically, a specific timepoint (such as 6 months) is chosen to evaluate whether a patient had an overall response or not. In addition to overall response, clinical benefit is another commonly used classification where a patient is considered to have a clinical benefit if they experience a CR, PR or an SD for 24 weeks or longer. Best overall response and clinical benefit can be modeled with ML using binary or multi-class classifiers.

For binary classification, we will use the Area Under the receiver operating characteristic (ROC) Curve, referred to as AUC, to evaluate our models’ prediction performance to correctly rank samples. Appropriate statistical methods will be used to find the confidence intervals (CIs) for the AUC values and for comparing different models in terms of AUC.^35^ For multi-class classification, we will use balanced accuracy, also called macro average recall.^36^ In addition, F1 score can be used to robustly evaluate model performance for unbalanced targets. Lower 95 CI of AUC > 0.5 and point estimate of AUC > TA will be defined as an acceptable success threshold for our single omics model to be combined in our multimodal data, where TA is a lower performance bound for the point estimate. We believe that TA=0.6 is a reasonable threshold based on existing research conducted in similar settings,^37^ however this threshold may be adjusted (i.e., stricter) such that a reasonable number of models are accepted.

### 5.3 Survival analysis

Progression-free survival (PFS) has been a widely accepted efficacy endpoint in oncology that reflects patients’ clinical outcome. It is defined as a time-to-event endpoint which is the time from randomization in a clinical trial to disease progression or death due to any cause. PFS will be the main outcome variable in survival analysis, although other endpoints such as overall survival, time to overall (tumor) response are also available in this project for subsequent investigation. These time-to-event endpoints can be modeled with ML using techniques such as random survival forests (RSF)^38^ and regularized Cox regression, and Cox proportional hazards deep neural networks.^39^ The Harrell’s concordance-index (C-index) will be used to evaluate models’ prediction performance in terms of assigning a higher risk to the patient that experiences an event first from any random pair of patients. Appropriate statistical methods will be used to find the CIs for C-index and for comparing different models in terms of C-index.^40^ Lower 95 CI of C-index > 0.5 and point estimate of C-index > TA will be defined as an acceptable success threshold for our single omics model to be combined in our multimodal data. We believe that TA=0.6 is a reasonable threshold based on existing research conducted in similar settings,^41^ however this threshold may be adjusted (i.e., stricter) such that a reasonable number of models are accepted.

### 5.4 Model fusion

Once two or more single models have passed their success criteria, we have several possible ways to combine them:^42,43^

- **Early fusion** (i.e, variables pooling): We combine the different modalities using their input covariates. The combined model will have new parameters for the covariates. This approach does not allow for side-by-side performance comparison between the single models but rather evaluating the information gained from the added input data.
- **Intermediate (joint) fusion** (i.e, embeddings): Another approach is to use intermediate outputs from single modality models (such as embeddings) to train a new model. An autoencoder could be fitted to learn a lower-dimensional data representation and we could use the encoder part as a feature extractor for new covariates.
- **Late fusion** (i.e, predictions pooling, ensemble model): We load pretrained models and combine their outputs. A naïve way is to average the predictions (weightless fusion) of the models. This approach allows us to precisely see how much a model contributes to the final prediction but may lack flexibility and thus yield subpar performance as compared to the early-stage approach. Similarly, we can train a new model (ensemble) using the predictions of each single model as input features. Using late-stage fusion allows us to quantify the respective contribution of each single model to the final prediction, using methods such as variable importance in a random forest ensemble model.

### 5.5 Model additive value

Our clinical model will be the baseline model for all comparisons, as it is the current gold standard. To show additive value from our omics model (either genomics and/or imaging) to our clinical model, we will investigate the null hypothesis *AUC*_clinical_ = *AUC*_clinical+omics_ for classification type endpoints and the null hypothesis *C* − *index*_clinical_ = *C* − *index*_clinical+omics_ for survival type endpoints. Thus, we are interested in showing that *AUC*_clinical_ ≤ *AUC*_clinical+omics_ or *C* − *index*_clinical_ ≤ *C* − *index*_clinical+omics_ using an appropriate statistical test. For the comparison of classification models (censoring not considered), one can use semi-parametric^44^ or non-parametric approaches.^35^ For the comparison of survival models, one can use the method proposed by Kang et al^40^ which compares two correlated C indices.

### 5.6 Model interpretation

Model interpretation is critical when exploring novel omics data to build trust in the model’s results. Some models are more interpretable than others due to their methods (i.e, decision tree, regression models) but may come at the cost of performance when compared to less interpretable models (i.e, deep learning). Although machine learning models can be difficult to interpret, various statistical and post-hoc methods exist to help with interpreting those otherwise black box models. To identify important prognostic factors, model agnostic methods such as Shapley values, feature importance scoring, and variations thereof can be applied for a wide range of machine learning model types. For images, several techniques can be used to understand important region of an image, either by masking important area and observing performance drops (occlusion technique) or by changing pixel values as a function of the loss (gradient based).^45^ Predictive factors can be viewed as interaction terms between the treatment assignment variable (binary, either 0 representing the control arm and 1 representing the investigational arm) and other baseline variables from any of the data modalities. Due to different data pre-processing and feature engineering techniques, any derived variable that involves the treatment assignment variable is considered as a predictive factor in the final model. Similar methods used to interpret prognostic factors apply here as well. Alternatively, methods that employ counterfactual outcome framework^46^ can also be used to identify predictive factors that lead to strongly differentiating subgroups in terms of treatment effect. Using this framework, prognostic models will firstly be used to impute the counterfactual outcome for each patient had they been on the other treatment arm, allowing the evaluation of treatment effect at individual patient level. A second-stage model is then built using the treatment effect as the outcome variable to identify the predictive factors.

### 5.7 Model training strategy

To robustly train our model using three data sources (**C**: clinical, **G**: genomics & transcriptomics and **I**: imaging), we designed rules that ensure safe and consistent comparisons between models during the investigation. Those rules are described below, and the training strategy is illustrated in **Figure** 3.

**Figure 3:**
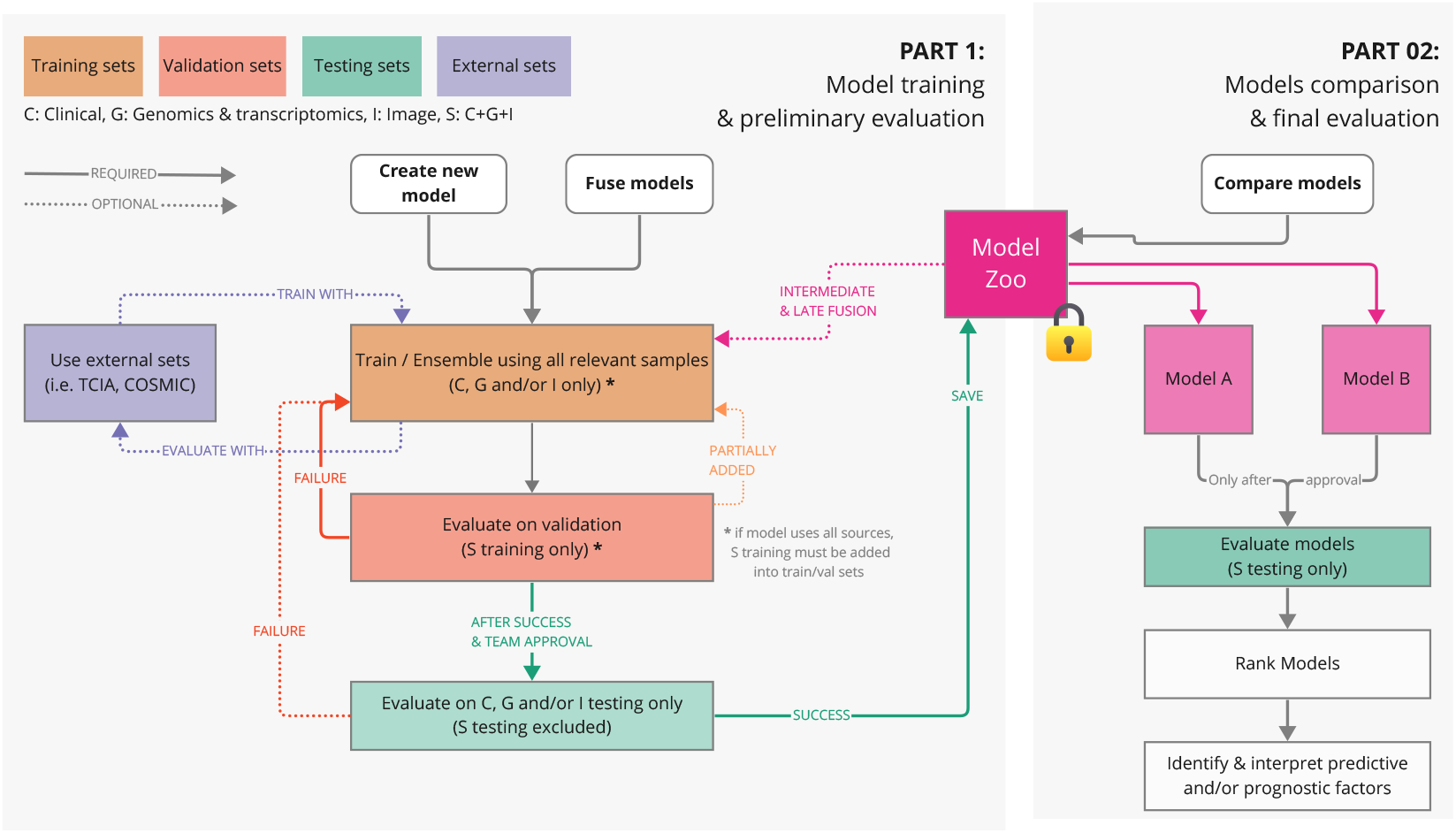
Overview of the required steps to train, fuse models and compare models. Successful model during the training part is saved into the model zoo. Comparison can include internal model as well as publicly available models,

1. **Training:** Each single (data source) model must be trained using all relevant (sample with available omics) samples from the training data, training_super_ excluded.
  - *OPTIONAL: External sets*: The use of external datasets is encouraged (see 2.2.2). Those data sets can increase our training size and/or allow model independent evaluation, both case helping our model generalize to unseen data and prevent overfitting.
  - *OPTIONAL: Pretrained model*: The use of pretrained models (from either our model zoo or public) is encouraged. Those models could be used for intermediate to late fusion (see 5.4) or simply for benchmarking (i.e, external gold standard model).
2. **Validation:** If the model is successful at Step #1 (based on 5.2 or 5.3), the model will be validated using the validation set (training_super_). This set contains all the data sources and thus allows us to consistently benchmark different -omics models with the same samples. If the model fails at this step, the user can modify the model and try again. We will not explicitly limit the number of times the validation set is accessed, but we will keep a log of the times validation set has been accessed to ensure that training_super_ has not been excessively reused. This validation will not constitute the final assessment of the model using clinical variables, as described in (6) below.
3. **Testing:** If the model is successful at Step #2 (based on 5.2 or 5.3), the next step is to validate on our test set (excluding testing_super_). This step requires approval from the extended team before using any testing sets so that the test set in not used repeatedly, because using the test set a few times may result in overfitting. If the model fails at this step, it will need to have significant design changes before it can be approved for testing with the test set again.
4. **Model zoo:** If the model is successful at Step #3, the model must be saved into our model zoo (repository to allow any users to load back the model, ensuring reproducibility for future use).
  - *OPTIONAL: Retrained model*: Before being saved to the model zoo, the user may train the model using training, validation sets used above to improve model performance and robustness to unseen data.
5. **Model comparison:** To compare models, we load them from our model zoo and evaluate them using the testing_super_ dataset. Each comparison can be done only once and only after team approval.

## 6 Summary

Novartis Pharmaceuticals Corporation and the FDA are collaborating on a research project to share and use multimodal data for developing ML models and finding prognostic and predictive factors for predicting clinical outcomes for HR+, HER2-metastatic breast cancer. This paper discussed data transfer procedures, internal and external datasets, data handling, pre-processing and splitting methods, proposed modeling strategies, endpoints, and success criteria for this collaboration. The internal data set, acquired through three randomized, double-blind, placebo-controlled, phase III clinical trials, provides rich data in terms of clinical, genomics and imaging information for modeling and discovery. Since our data set is retrospectively adapted for use in modeling, it presents challenges for modality availability, missingness, filtering, data splitting, and model fusion. The statistical and ML analysis plans presented in this paper provide a roadmap through which we intend to design various models for this rich and complex data set and to evaluate their performance.

## Data Availability

All data for the MONALEESA studies are available at clinicaltrials.gov and Kisqali USPI.

## 7 Acknowledgments

This work was supported in part through an Office of Women’s Health grant from the U.S. Food and Drug Administration, and by an appointment to the Research Participation Program at the Center for Devices and Radiological Health administered by the Oak Ridge Institute for Science and Education through an interagency agreement between the U.S. Department of Energy and the U.S. Food and Drug Administration.

### 8 Appendix

#### 8.1 Model training example

1. Here is a practical example of **training a model from scratch** using any fusion approach:
  a. Our training data set will consist of the data in all training sets except training_super_ (i.e, training_clinical_, training_genomics_, training_imaging_. It allows the user to freely decide how to train and perform initial evaluation of the model (cross-validation, leave-one-out, etc.) while minimizing the need for validation and test datasets. The reason these three datasets (training_clinical_, training_genomics_, training_imaging_ can be combined for training a clinical model is that baseline clinical variables are contained in each of them, as discussed in “Data handling and pre-processing” Section.
    - *OPTIONAL: External sets*: The use of external datasets is encouraged when possible (see 2.2.2). Those datasets can increase our training size or evaluate our model using independent data. In both cases it would help our model generalize to unseen data and prevent overfitting.
    - *OPTIONAL: Pretrained model*: The use of pretrained models (from either our model zoo or public) is encouraged when applicable. Those models could be used for intermediate to late fusion (see 5.4) or simply for benchmarking (i.e, external model being the gold standard).
  b. If the model is successful at Step #1, the model will be validated using the validation set (training_super_).
  c. If the model is successful at Step #2, the next step is to validate on our test set (excluding testing_super_). To test a clinical model, the user could use all available clinical information (i.e. testing_clinical_, testing_genomics_, testing_imaging_). The reason these three datasets can be combined for testing the model is that baseline clinical variables are contained in each of them, as discussed in “Data handling and pre-processing” Section.
  d. If the model is successful at Step #3, the model must be saved into our model zoo.
  e. The final assessment of the model with clinical variables saved into the model zoo will be performed with the testing_super_ data set, as described in (D) below.
2. Here’s a practical example of **estimating model performance**:
  a. We load the model (e.g., either model_clinical_ or model_genomics_) from our model zoo
  b. The model is evaluated on the testing_super_ + relevant testing sets. We use the results to quantify and interpret the performance.
3. Here is a practical example of **comparing two models** from our model zoo:
  a. We load the two models (model_clinical_ and model_genomics_) from our model zoo.
  b. The two models are evaluated on the testing_super_.
  c. We use the results to quantify, rank and compare the performances.
4. Here is a practical example of **training a super model** (genomics + imaging) using any fusion method, as seen on **Figure** 4:

**Figure 4:**
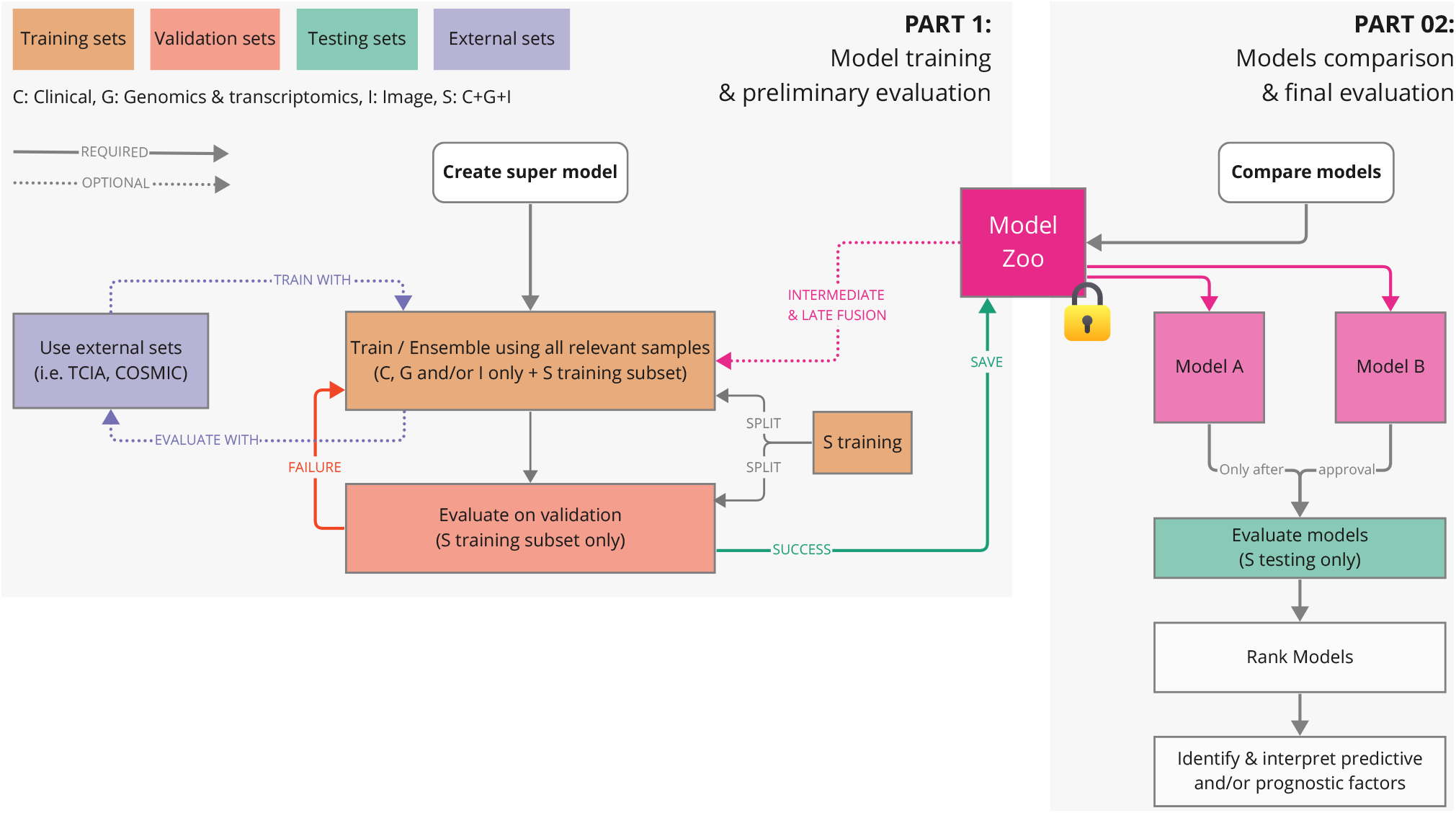
Example of an early fusion model of super samples. updated model strategy for super training model
  a. Our training_super_ is divided into two subsets. One is added to our training samples and the other stays as our internal validation.
  b. *Optional* For intermediate and late fusion, we load our model(s) from the zoo.
  c. Similarly, as seen in examples above, we can train and validate our model following the established guidelines.
  d. If the model has a successful validation, it will be saved in our model zoo.
  e. Model comparisons are same as for any other models, either standalone or to compare against others.

## Notes

### Competing Interest Statement

The authors have declared no competing interest.

### Clinical Trial

NCT01958021, NCT02422615, NCT02278120

### Clinical Protocols

https://clinicaltrials.gov/study/NCT01958021?intr=kisqali&rank=4

https://clinicaltrials.gov/study/NCT02422615?term=monaleesa-3&intr=Ribociclib%20%5C(LEE011%5C)&rank=1

https://clinicaltrials.gov/study/NCT02278120?term=monaleesa-7&intr=Ribociclib%20%5C(LEE011%5C)&rank=1

### Funding Statement

This study did not receive any funding

### Author Declarations

Ethics Committee from Novartis gave ethical approval for the MONALEESA trials

